# Longitudinal changes in cannabis use and misuse in the 5 years following recreational cannabis legalization in Canada: A prospective cohort study of community adults

**DOI:** 10.1101/2024.08.06.24311571

**Authors:** André J. McDonald, Amanda Doggett, Kyla Belisario, Jessica Gillard, Jane De Jesus, Emily Vandehei, Laura Lee, Jillian Halladay, James MacKillop

**Author notes:** **Corresponding author** André McDonald Peter Boris Centre for Addictions Research, McMaster University 100 West 5th Street, Hamilton, Ontario, Canada, L8P 3P2.

## Abstract

**Importance:** A growing number of jurisdictions have legalized recreational cannabis for adults, but most evaluations have used repeated cross-sectional designs, preventing examination of within-person and subgroup trajectories across legalization.

**Objective:** To examine changes in cannabis use and misuse in the five years following legalization in Canada – the first G7 country to legalize adult recreational cannabis use – both overall and by pre-legalization cannabis use frequency using a longitudinal design.

**Design:** Prospective cohort study with 11 biannual assessments from September 2018 to October 2023. Mean retention was 90% across all waves.

**Setting:** Ontario, Canada.

**Participants:** Sample of 1,428 (60.2% female, M_age_=34.5) community adults aged 18 to 65 years.

**Exposure:** Five years of recreational cannabis legalization (the baseline wave was immediately prior to legalization).

**Main outcome and measures:** Primary outcomes were cannabis use frequency and cannabis misuse (CUDIT-R score). Pre-legalization cannabis use frequency, age, and sex were examined as moderators. Secondary outcomes included changes in cannabis product preferences over time.

**Results:** Linear mixed effects modelling found a significant increase in cannabis use frequency, such that the mean proportion of days using cannabis increased by 0.35% (*p*<.001) per year in the overall sample (1.75% over 5 years). In contrast, CUDIT-R scores (on scale of 0 to 32) decreased significantly overall (b=-0.08 [-0.4 over 5 years], *p*<.001), most notably following the onset of the COVID-19 pandemic. Interaction analyses indicated that pre-legalization cannabis use frequency significantly moderated changes for both outcomes (*p*<.001). Specifically, cannabis use and misuse decreased among pre-legalization frequent consumers and modestly increased among occasional/non-users. Cannabis product preferences shifted away from dried flower, hashish, concentrates, oil, tinctures, and topicals to edibles, liquids, and vape pens.

**Conclusions and Relevance:** In the five years following legalization, cannabis use frequency increased modestly, while cannabis misuse decreased modestly in a longitudinal observational cohort of Canadian adults. These changes were substantially moderated by pre-legalization cannabis use, with more frequent consumers of cannabis pre-legalization exhibiting the largest decreases in both outcomes. Although longer-term surveillance is required, these results suggest Canadian recreational cannabis legalization was associated with modest negative consequences and some evidence of positive outcomes among nonclinical community adults.

**Key points:** *Question:* Did cannabis use or misuse change among adults in the five years following recreational cannabis legalization in Canada (overall and by pre-legalization cannabis use frequency)?

*Findings:* Overall, cannabis use frequency increased significantly while misuse decreased significantly, with small effect sizes for both. Pre-legalization cannabis use significantly moderated these changes. Product preferences shifted from dried flower, hashish, and concentrates to edibles, liquids, and vape pens.

*Meaning:* From a public health standpoint, these findings suggest both a modest negative consequence (small increase in cannabis use frequency) and positive outcomes (small decrease in cannabis misuse, and transition from combustible to non-combustible products).

## Introduction

A growing number of jurisdictions have liberalized, or are considering liberalizing, the use of recreational (non-medical) cannabis. In October 2018, Canada became the first G7 country to legalize recreational cannabis use for adults, serving as an experiment for other countries considering national policy reform. Leading up to legalization in Canada, there were concerns that cannabis use and misuse would increase due to easier access, growing social acceptability, declining perception of harm, product diversification, and increasing potency.^1–4^ Since legalization, some evidence indicates that these concerns were justified,^5,6^ although other investigations have identified only limited negative consequences.^7,8^

In general, previous research evaluating the impact of recreational cannabis legalization on cannabis use and misuse has found mixed results. For example a repeated cross-sectional survey in the US found that states that legalized recreational cannabis use saw significant post-legalization increases in the prevalence of cannabis use, frequent cannabis use, and cannabis use disorders among adults aged 26 years and older, but no significant changes among young adults aged 18 to 25 years.^9^ However, another repeated cross-sectional study found that increasing cannabis use in the US was due to general period effects and not legalization.^8^ In Canada, repeated cross-sectional studies predominantly suggest that legalization has been associated with increases in the prevalence of cannabis use and misuse among adults.^5,10–12^ Healthcare utilization studies have also suggested that cannabis-related emergency department visits and hospitalizations have increased including cannabis use disorders, poisonings from edibles, cannabis-induced psychosis, self-harm involving cannabis, and other cannabis-attributable conditions.^13–20^

An important limitation of the cannabis legalization literature is that most previous studies have used a repeated cross-sectional design, which does not allow for the examination of within-person changes from pre-legalization to post-legalization. Very few within-person studies have evaluated the impact of legalization on cannabis use and misuse, representing a significant research gap.^21,22^ Longitudinal designs are necessary to evaluate within-person changes, as well as subgroup trajectories across legalization such as pre-legalization cannabis use levels and sex. Another significant limitation of the current literature is that most studies focus on the early stages of legalization; it is widely acknowledged that understanding the impacts of legalization requires a longer post-legalization time frame.^7,23,24^

Here we report changes in cannabis use and misuse over the five years following legalization in a nonclinical community-based cohort of Canadian adults, with repeated measures collected approximately every six months. The original wave of data collection took place during the month prior to legalization and an initial report on the 12-month outcomes found small increases in use and misuse.^22^ The current study’s first objective was to examine changes in cannabis use and misuse over a full five years since recreational legalization in Canada. The second objective was to examine whether pre-legalization cannabis use frequency moderated these changes – that is, whether changes depended on how frequently one used cannabis pre-legalization. This permitted evaluating whether exacerbations were present among already frequent consumers. The third objective was to examine whether cannabis product type preferences among active users changed over the five years since legalization.

## Methods

### Cohort and Study Design

Participants were recruited from an existing registry of ambulatory community adults at St. Joseph’s Healthcare Hamilton in Hamilton, Ontario, Canada. Eligibility criteria included being between the ages 18 and 65 years at baseline, minimum ninth-grade education, willingness to consider participation in research studies, and no extant terminal illness. Registry enrollment involved a single extended in-person assessment.

To create the cannabis legalization surveillance cohort, we invited registry members to enroll in a supplementary study comprising periodic online assessments, with the baseline occurring in the month prior to the legalization of recreational cannabis for adults on October 17, 2018 (from mid-September to mid-October 2018). Participants were required to accept the invitation and provide informed consent (n=1502). Subsequent waves of data collection were conducted each April and October up to October 2023 (11 waves total). Participants received an online gift card ($40 CAD) upon completion of each wave. All procedures were approved by the Hamilton Integrated Research Ethics Board (Project #4699). Data quality was protected using questions with unambiguously correct responses (e.g., “*For this item, select option B*”), with participants required to answer 3+ correctly for a given wave. Participants with less than 3 total follow-up observations were excluded, producing a final sample size of n=1,428 (95% overall inclusion). The cohort had a high retention rate across waves 2 to 11 (95.0%, 93.1%, 93.4%, 93.0%, 91.3%, 88.9%, 88.1%, 86.9%, 86.3%, 86.5%; mean retention rate=90.2%; see eFigure 1 for flowchart of exclusions and loss to follow up). Missing data strategies are presented in the *Statistical Analyses* section.

Table 1 shows the baseline characteristics of the final study cohort. The cohort was slightly overrepresented by females (60.2%), predominantly white (78.9%) and unmarried (68.3%), highly educated (92.0% with at least some post-secondary), had an average age of 35 years and a median income of $45,000 to $90,000. Roughly half the sample reported no cannabis use, and half reported some use with 16% scoring 6 or higher on the CUDIT-R indicating probable cannabis misuse.^25^

**Table 1:** Baseline (pre-legalization in September 2018) characteristics of the final study cohort (N=1,428). A total CUDIT-R score of 6+ is used as a cut-off to indicate probable cannabis misuse.^25^

| Indicator | n (%) |
| --- | --- |
| Sex |  |
| Male | 569 (39.8%) |
| Female | 859 (60.2%) |
| Age (*Mean (SD)*) | 34.5 (13.9) |
| Race |  |
| White | 1,127 (78.9%) |
| Non-white | 301 (21.1%) |
| Household income |  |
| Less than \$45,000 | 442 (31.0%) |
| \$45,000 to \$90,000 | 437 (30.6%) |
| \$90,000+ | 549 (38.4%) |
| Education |  |
| High school or less | 114 (8.0%) |
| Some post-secondary | 661 (46.3%) |
| Bachelor's degree | 495 (34.7%) |
| Postgraduate/professional degree | 158 (11.1%) |
| Marital status |  |
| Unmarried | 975 (68.3%) |
| Married | 453 (31.7%) |
| Cannabis use frequency |  |
| None | 749 (52.4%) |
| <Monthly | 246 (17.2%) |
| Monthly | 168 (11.8%) |
| Weekly | 141 (9.9%) |
| Daily+ | 124 (8.7%) |
| CUDIT-R score |  |
| No cannabis use | 749 (52.5%) |
| <6 | 450 (31.5%) |
| 6+ | 229 (16.0%) |

### Measures

Two primary outcomes were examined – cannabis use frequency and cannabis misuse. Cannabis use frequency was measured with the following question from the Canadian Cannabis Survey^12^: *In the past 6 months, how often did you typically use cannabis? Never, <1 day/month, 1 day/month, 2−3 days/month, 1–2 days/week, 3–4 days/week, 5–6 days/week, or Daily*. These categories were transformed into continuous values representing the proportion of days using cannabis; for categories with a range, we used the mid-point (i.e., Less than one day per month = 0.5/30.435 = 0.016; One day per month = 1/30.4347 = 0.033; 2 to 3 days per month = 2.5/30.435 = 0.082; 1 or 2 days per week = 1.5/7 = 0.214; 3 or 4 days per week = 3.5/7 = 0.5; 5 to 6 days per week = 5.5/7 = 0.786; Daily = 1). These values were then multiplied by 100 so that beta coefficients would represent the percentage of days using cannabis for ease of interpretation. Cannabis misuse was measured with the 8-item CUDIT-R.^26^ Responses were summed to generate a total CUDIT-R score, providing a continuous measure of cannabis misuse that ranged from 0 to 32. Both outcomes were treated as continuous variables.

The secondary set of 11 outcomes included reporting use of various cannabis products including: dried flower/leaf (smoked or vaporized), hashish, cannabis oil from a Health Canada licensed producer, liquid concentrate (e.g., hash oil, butane honey oil, etc.), cannabis oil cartridges or disposable vape pens, solid concentrate (e.g., shatter, budder, etc.), edibles (e.g., prepared food products), liquid (e.g., cola/tea), tinctures (e.g., concentrated amounts ingested orally or taken under the tongue), topical ointments (e.g., lotions, salves, balms applied directly to the skin), and fresh flower/leaf (e.g., for juicing). Each cannabis product outcome was treated as binary (yes or no). This list of cannabis products were based measures from the Canadian Cannabis Survey.^12^

Covariates included baseline age, sex assigned at birth (male or female), ethnicity (white or non-white), marital status (unmarried or married), household income (less than $45,000; $45,000 to $90,000; $90,000+), and education (high school or less; some post-secondary; bachelor’s degree; postgraduate/professional degree). Pre-legalization cannabis use frequency (never, <monthly, monthly, weekly, daily+) was examined as a moderator.

### Statistical Analyses

In our primary analyses, we conducted linear mixed-effects models to examine trends over time in cannabis use frequency (proportion of days using cannabis) and cannabis misuse (CUDIT-R score), controlling for baseline age, sex, ethnicity, marital status, household income, education, and pre-legalization cannabis use frequency. We also explored pre-legalization (baseline) cannabis use frequency, age, and sex as moderators for the association between time (continuous and rescaled to yearly changes) and the two outcomes. For each outcome, we initially included the identified covariates and the main effect of time only, then tested a two-way interaction between time and pre-legalization cannabis use frequency in a separate model.

We performed an attrition analysis and found that completing less than 2 follow-ups (those excluded) was significantly related to baseline cannabis use frequency and income, but no other study variables (see eTable 1). To explore the potential impact of attrition bias, we performed a sensitivity analysis by modelling the outcomes using a joint analysis and Bayesian imputation framework through the R package JointAI.^27^ Although omnibus tests were not available from the JointAI models, coefficient estimates were consistent with the main analysis suggesting missing data did not bias the results significantly (see eTable 2).

In our secondary analyses, we examined changes in cannabis product preferences among active cannabis users at each wave (i.e., only participants’ observations reporting cannabis use) by estimating adjusted prevalence differences using multivariable modified least-squares regression models.^28^ Time (continuous) was the predictor and reporting use of a given cannabis product (yes or no) was the binary outcome. We used R version 4.1.0 to conduct all statistical analyses and create data visualizations, and reported our findings in accordance with STROBE guidelines.^29^ *P*<.05 and 95% confidence intervals that did not include the null were considered statistically significant.

## Results

### Overall changes in cannabis use and moderation by pre-legalization cannabis use frequency

Main effects of time and interactions are in Table 2. The main effect model found a significant increase in cannabis use frequency in the overall sample (b=0.35; p<0.001), such that the mean proportion of days using cannabis increased by 0.35% per year (i.e., over 5 years, a 1.75% increase). Figure 1A shows longitudinal changes in cannabis use frequency since legalization in the overall sample.

**Table 2:** Main effect of time and interaction effect of pre-legalization cannabis use frequency for cannabis use frequency and CUDIT-R.

|  | Outcome 1: Cannabis use frequency |  |  |  | Outcome 2: CUDIT-R |  |  |  |
| --- | --- | --- | --- | --- | --- | --- | --- | --- |
|  |  |  | Omnibus test |  |  |  | Omnibus test |  |
| | $\beta$ | $SE$ | $F$ | p | $\beta$ | $SE$ | $F$ | p |
|  | *Main effect models* |  |  |  |  |  |  |  |
| Time (annual changes) | 0.35 | 0.08 | **18.71** | **<0.001** | -0.08 | 0.01 | **53.41** | **<0.001** |
|  | *Interaction models* |  |  |  |  |  |  |  |
| Time (annual changes) | 0.82 | 0.11 | **19.95** | **<0.001** | 0.10 | 0.01 | **379.87** | **<0.001** |
| Pre-legalization cannabis use |  |  | **773.14** | **<0.001** |  |  | **531.09** | **<0.001** |
| None | Ref | - |  |  | Ref | - |  |  |
| Less than monthly | 1.74 | 1.31 |  |  | 1.50 | 0.18 |  |  |
| Monthly | 9.28 | 1.53 |  |  | 3.18 | 0.21 |  |  |
| Weekly | 36.74 | 1.64 |  |  | 6.18 | 0.23 |  |  |
| Daily | 91.01 | 1.74 |  |  | 9.95 | 0.24 |  |  |
| Time* Pre-legalization cannabis use |  |  | **121.32** | **<0.001** |  |  | **130.71** | **<0.001** |
| Time*Less than monthly | 0.40 | 0.22 |  |  | -0.21 | 0.03 |  |  |
| Time*Monthly | 0.50 | 0.25 |  |  | -0.32 | 0.03 |  |  |
| Time*Weekly | -1.14 | 0.28 |  |  | -0.50 | 0.04 |  |  |
| Time* Daily | -6.08 | 0.29 |  |  | -0.73 | 0.04 |  |  |

Notes: All models adjusted for baseline age, sex, ethnicity, income, education, and marital status.

**Figure 1.**
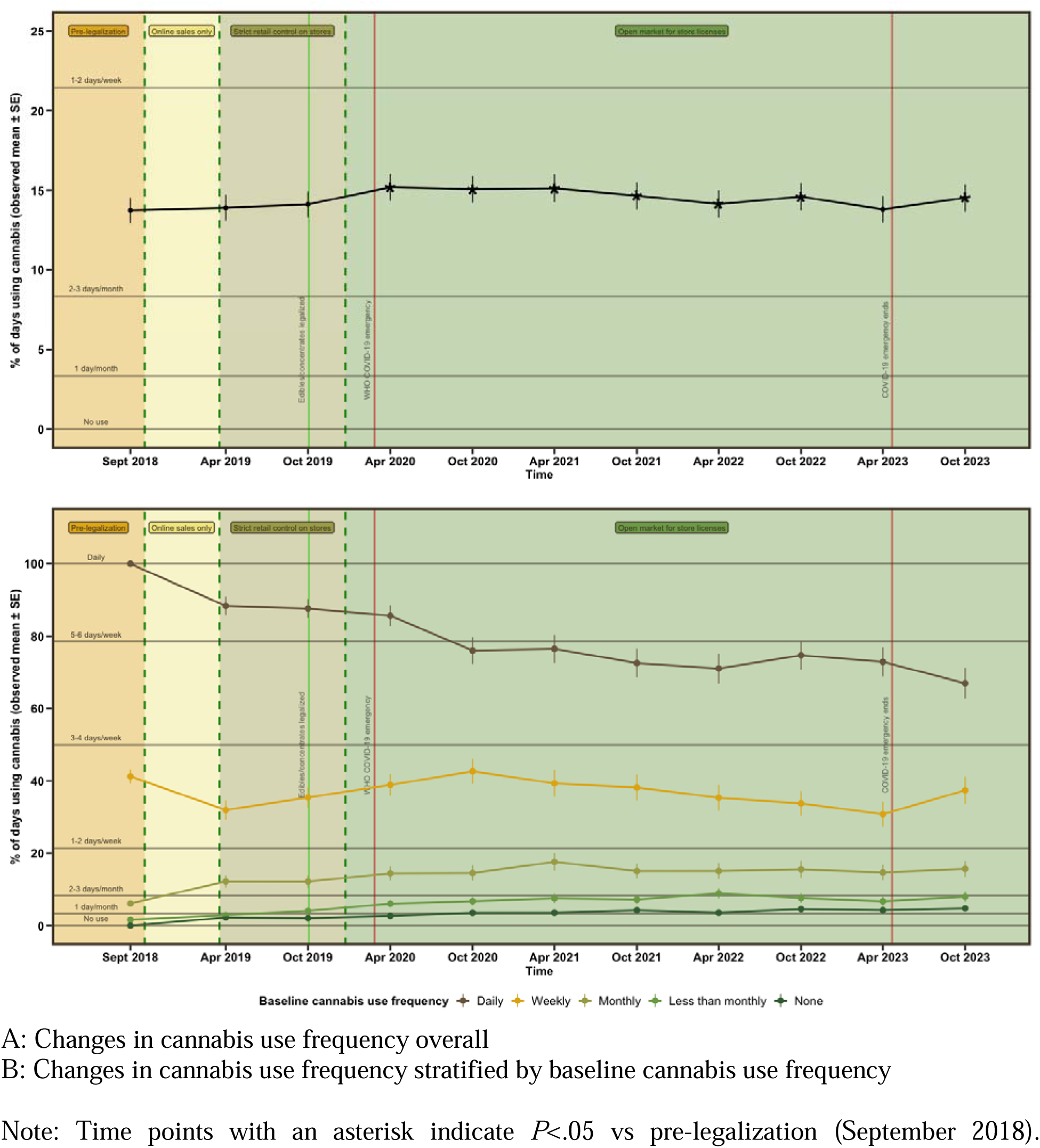
Mean (±SE) for cannabis use frequency over 5 years since legalization from September 2018 to October 2023 (10 waves) in the overall sample and stratified by baseline cannabis use frequency.

As shown in Table 2, we found a significant interaction between time and pre-legalization cannabis use frequency (*p*<0.001). Figure 1B shows longitudinal changes in cannabis use frequency since legalization, stratified by pre-legalization cannabis use frequency. We observed a significant decrease among those using daily at baseline, little change for those using weekly at baseline, and slight increases among those using monthly or less at baseline. To qualitatively characterize the patterns of change, Figure 2 provides a person-centred alluvial plot showing transitions between cannabis use frequency groups from the first wave (pre-legalization in September 2018) to the last wave (5 years post-legalization October 2023). Wave-by-wave transitions are shown in eFigure 2A.

**Figure 2:**
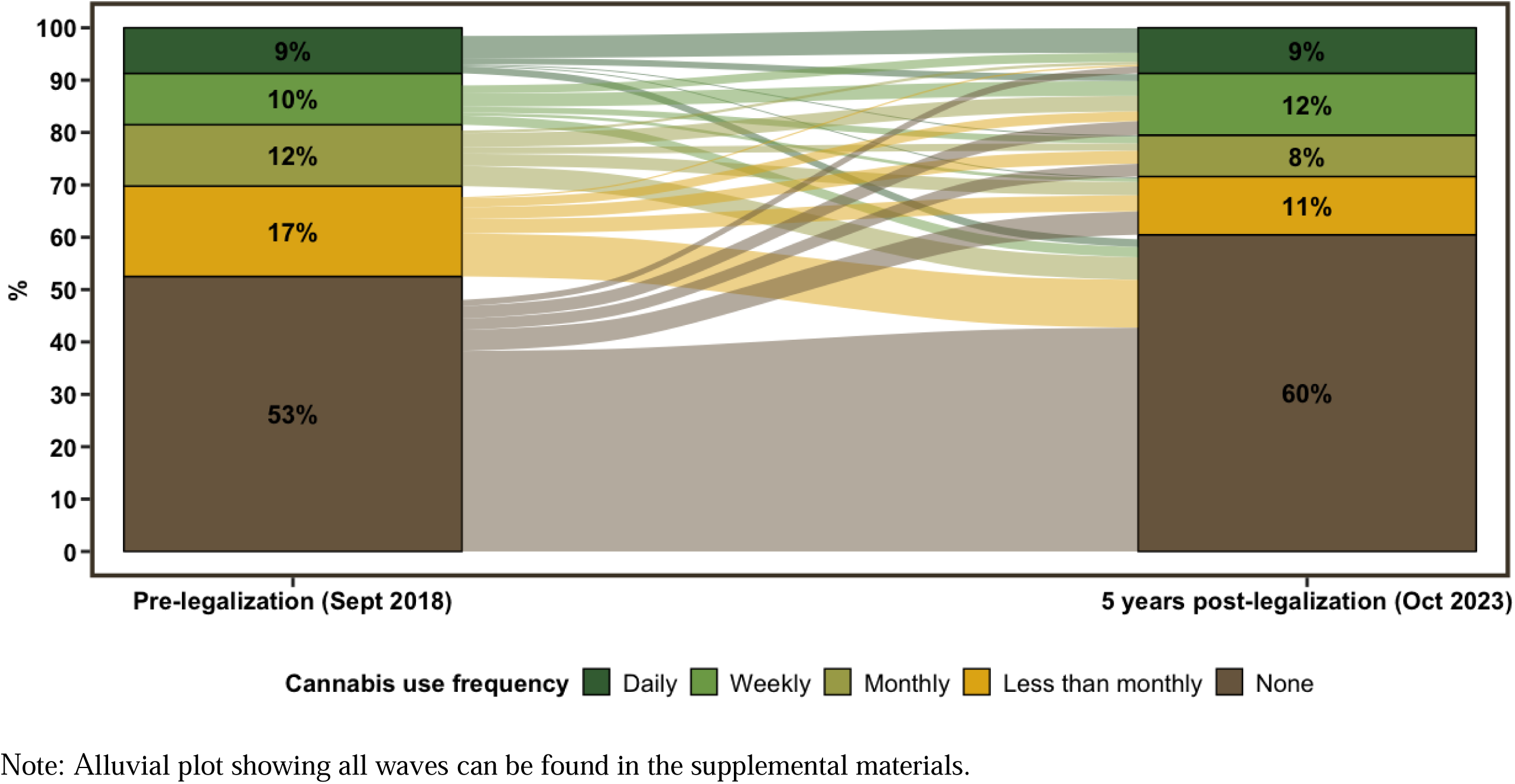
Alluvial plots showing transitions in cannabis use frequency from pre-legalization (September 2018) to 5 years post-legalization (October 2023).

### Overall changes in cannabis misuse and moderation by pre-legalization cannabis use frequency

Main effects of time and interactions are presented in Table 2. The main effect model found a significant decrease in CUDIT-R score in the overall sample (b =-0.08; *p*<0.001), such that the mean CUDIT-R score decreased by 0.08 points per year (i.e., over 5 years, a 0.4 point decrease on average). In a follow-up analysis that removed the first item from the CUDIT-R outcome (cannabis use frequency), we found a similar result (b =-0.07; *p*<0.001). Figure 3A shows longitudinal changes in CUDIT-R score since legalization in the overall sample. Interestingly, we observed a notable decrease in cannabis misuse that occurred during the early phase of the COVID-19 pandemic (April to October 2020).

**Figure 3.**
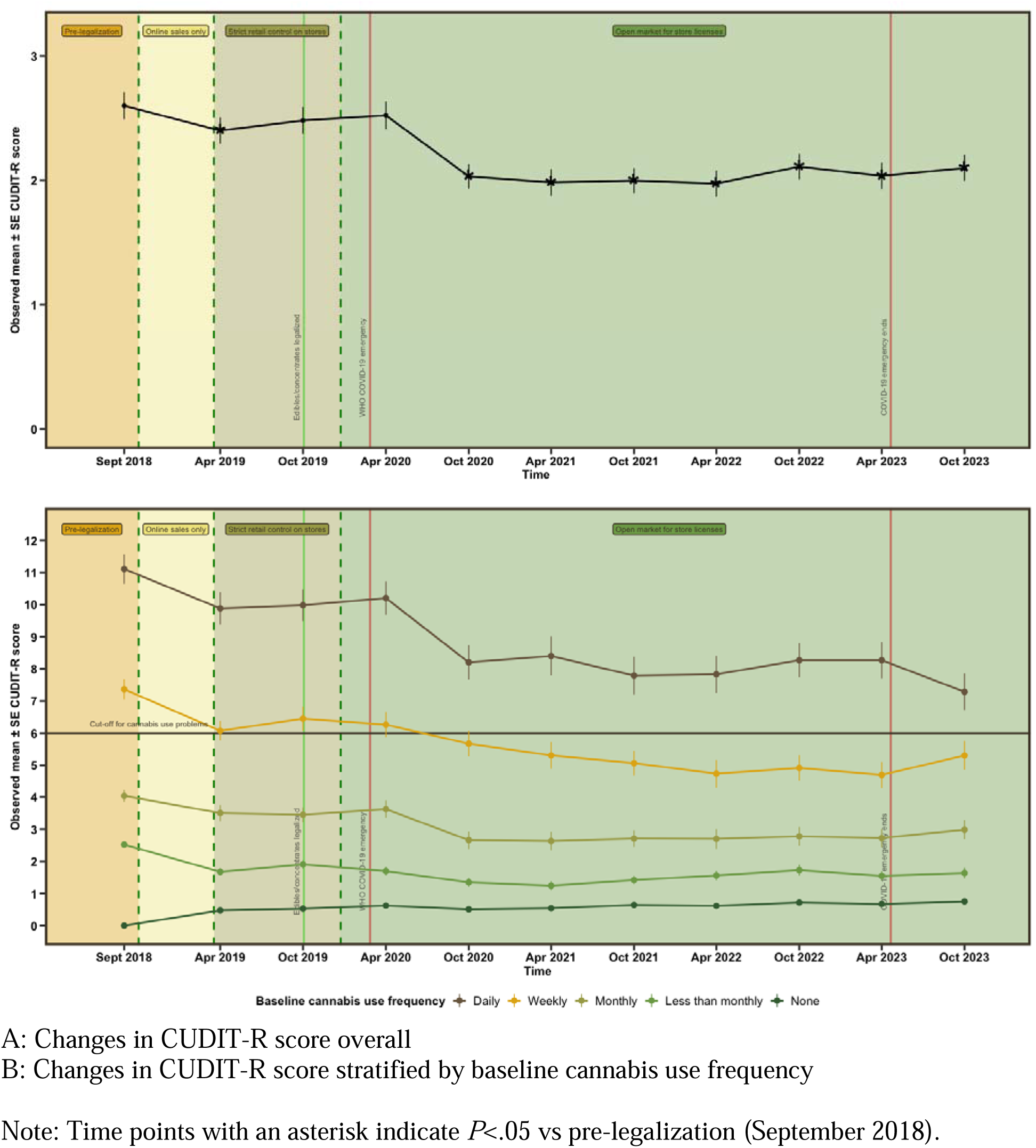
Mean (±SEM) for CUDIT-R score over 5 years since legalization from September 2018 to October 2023 (10 waves) in the overall sample and stratified by baseline cannabis use frequency.

As shown in Table 2, we found a significant interaction between time and pre-legalization cannabis use frequency (*p*<0.001). Figure 3B shows longitudinal changes in cannabis use frequency since legalization stratified by pre-legalization cannabis use frequency. We observed a significant decrease among those using monthly or less than monthly at baseline, and a slight increase among those not using at baseline. Of note, those using weekly at baseline, on average, crossed from above to below the validated CUDIT-R cut-off score of 6 indicating probable cannabis misuse.^25^ To qualitatively characterize the patterns of change, Figure 4 provides a person-centred alluvial plot showing transitions between cannabis misuse groups from the first wave (pre-legalization in September 2018) to the last wave (5 years post-legalization October 2023). Wave-by-wave transitions are shown in eFigure 2B.

**Figure 4:**
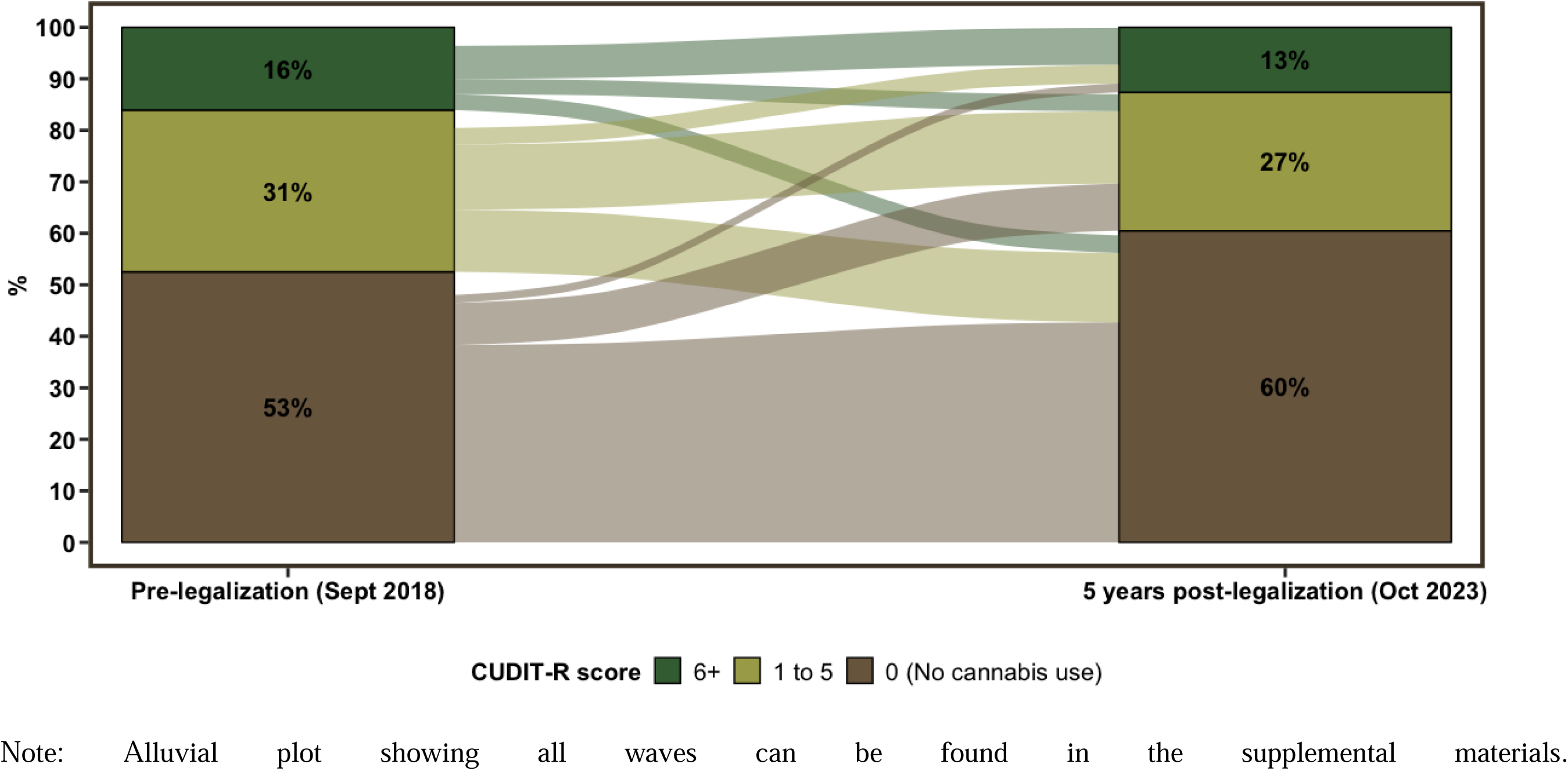
Alluvial plots showing transitions in CUDIT-R scores from pre-legalization (September 2018) to 5 years post-legalization (October 2023).

### Changes in cannabis product preferences

Table 3 presents results from our secondary analyses estimating adjusted prevalence differences (annual percentage changes) in cannabis product preferences among active cannabis users. We observed statistically significant decreases in dried flower, solid concentrate, liquid concentrates, cannabis oil, tinctures, topical ointments, and hashish. Conversely, we observed statistically significant increases in edibles, liquids, and cannabis oil cartridges or disposable vape pens. The most pronounced decrease was in dried flower, with a 3.56% (95% CI: 2.91%, 4.22%) annual decrease in prevalence among active cannabis users (from 81.3% pre-legalization to 64.6% at 5 years post-legalization). The most pronounced increase was in cannabis oil cartridges or disposable vape pens, with a 3.39% (95% CI: 2.72, 4.05%) annual increase in prevalence among active cannabis users (from 18.4% pre-legalization to 33.0% at 5 years post-legalization).

**Table 3:** Adjusted prevalence differences (annual percentage changes) in cannabis product preferences among active cannabis users in the 5 years since legalization estimated from multivariable modified least squares regression models.

| Cannabis product use (outcome) | Time in years (predictor) |  |  | p | aPD x 5 years |
| --- | --- | --- | --- | --- | --- |
|  | aPD | 95% CI |  |  |  |
| Dried flower/leaf (smoked or vaporized) | -3.56% | -4.22% | -2.91% | <0.001 | -17.80% |
| Solid concentrate (e.g., shatter, budder, etc.) | -2.22% | -2.68% | -1.76% | <0.001 | -11.10% |
| Liquid concentrate (e.g., hash oil, butane honey oil, etc.) | -1.45% | -1.88% | -1.03% | <0.001 | -7.25% |
| Cannabis Oil from a Health Canada Licensed Producer | -0.95% | -1.53% | -0.38% | 0.001 | -4.75% |
| Tinctures (e.g., concentrated amounts ingested orally or taken under the tongue) | -0.89% | -1.34% | -0.45% | <0.001 | -4.45% |
| Topical Ointments (e.g., lotions, salves, balms applied directly to the skin) | -0.79% | -1.27% | -0.30% | 0.001 | -3.95% |
| Hashish | -0.72% | -1.15% | -0.29% | 0.001 | -3.60% |
| Fresh flower/leaf (e.g., for juicing) | -0.08% | -0.24% | 0.07% | 0.29 | -0.40% |
| Edibles (e.g., prepared food products) | 1.22% | 0.46% | 1.99% | 0.002 | 6.10% |
| Liquid (e.g., cola/tea) | 2.21% | 1.77% | 2.66% | <0.001 | 11.05% |
| Cannabis oil cartridges or disposable vape pens | 3.39% | 2.72% | 4.05% | <0.001 | 16.95% |

Notes: aPD = adjusted prevalence difference; 95% CI = 95% confidence interval. All models adjusted for baseline age, sex, ethnicity, income, education, and marital status, and used robust variance estimation.

Figure 5 presents the proportion of active cannabis users reporting use of dried flower, oil cartridges/vape pens, edibles, liquids, and solid concentrate (the most dynamic products) over time since legalization.

**Figure 5:**
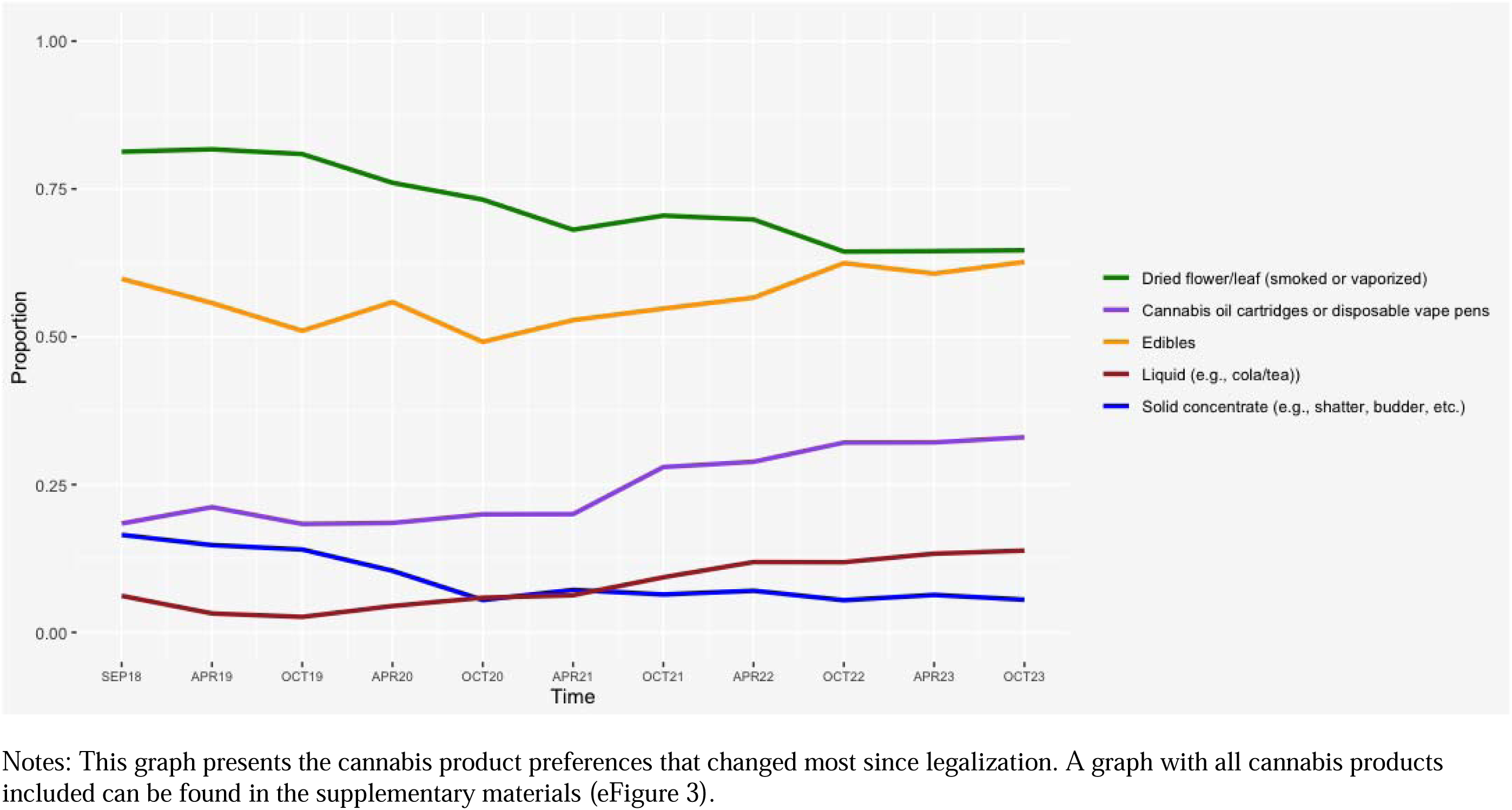
Cannabis product preferences over time since cannabis legalization among active cannabis users at each wave.

### Sex and age moderation of changes over time

We examined sex and age as moderators for change in cannabis use frequency and CUDIT-R score over time. When testing age as a moderator, we dichotomized age to separate young adults (under 30 years of age), who typically age out of cannabis use,^30^ from middle-aged and older adults (30+ years of age). No interactions were present for frequency, but interactions were present for sex and age in relation to CUDIT-R (see eTable 3 and eFigure 4), such that reductions were more pronounced among males and younger participants (particularly the latter), consistent with higher CUDIT-R scores in males and younger participants.

## Discussion

This study found that cannabis use frequency modestly increased while cannabis misuse decreased over the first 5 years of legalization in a nonclinical cohort of adults. From a public health standpoint, these results are mixed as increased use might be considered harmful, while decreased misuse is a positive outcome. Given that age was a significant moderator, whereby younger adults had larger declines in problems, these mixed findings may partly be due to the ‘aging out’ developmental trajectory that typically occurs during young adulthood,^30^ which appeared to affect cannabis misuse more than cannabis use frequency. Similar results have been observed in another cohort study,^21^ which found a significant decrease in cannabis-related adverse consequences but little change in cannabis use frequency.

Cannabis misuse notably declined immediately after the onset of the COVID-19 pandemic, and never returned to pre-pandemic levels. This was most pronounced among pre-legalization high-frequency consumers. We note that another longitudinal study similarly found that among pre-COVID frequent cannabis users, cannabis use frequency increased early in the pandemic while cannabis use disorder symptom severity decreased slightly.^31^ Understanding the environmental or psychological factors leading to these changes warrants further investigation.

When examining pre-legalization cannabis use as a moderator for the association between time and cannabis use frequency, we found a significant interaction such that cannabis use frequency decreased among those already using frequently pre-legalization and increased among pre-legalization abstainers. Changes in cannabis misuse were similarly moderated by pre-legalization cannabis use frequency, such that cannabis misuse decreased for all groups already using cannabis pre-legalization and increased for pre-legalization non-users. These findings do not support the notion that cannabis legalization would amplify or otherwise exacerbate existing patterns of use among active consumers. For both outcomes, it is also possible that regression to the mean explains part of the interaction effects. Fundamentally, however, these results do not suggest increased adverse outcomes for adults who were actively using cannabis pre-legalization.

Secondary analyses revealed significant changes in cannabis product preferences among those using cannabis actively over time, with decreases in dried flower, concentrates, cannabis oil, tinctures, topical ointments, and hashish, and increases in edibles, liquids, and cannabis oil cartridges/disposable vape pens. Our findings are similar to those of the International Cannabis Policy Study, which also found a pronounced decrease in dried flower, and increases in edibles, oils and drinks.^5^ We note that edibles and liquids only became legal in Canada on October 17, 2019, one year after cannabis flower and oils became legal.^32^ From a lung health perspective, it is a positive development that cannabis users transitioned away from dried flower, which is typically combusted, and towards non-combusted oils, edibles, and drinks. However, it is potentially concerning that cannabis drinkables are becoming more popular given how little research has examined their health effects.

### Strengths and limitations

This study addresses important research gaps and adds a new dimension to the small but growing cannabis legalization evaluation literature, complementing healthcare utilization studies and repeated cross-sectional surveys, which have mostly found negative outcomes associated with legalization for adults.^10–20^ To our knowledge, it uses the longest follow-up of any within-subjects longitudinal study evaluating recreational cannabis legalization, providing a more nuanced understanding of cannabis behaviour changes following legalization. Our prospective cohort had a high retention rate, averaging 90% retention across all follow-up waves, with 87% remaining at the five-year mark. The study also had limitations. Without comparison to a jurisdiction without cannabis legalization, it is difficult to attribute changes in cannabis use and misuse to legalization per se. It is possible that age-related decreases partly explained the interaction effects we observed. We also only had one pre-legalization time point and were therefore unable to conduct an age-period-cohort analysis. Considering the cohort was recruited from a community-level research registry, the results may not be fully generalizable nationally or internationally, especially given the diversity of policies and regulations accompanying cannabis legalization in other jurisdictions.

### Conclusions

This study found a small increase in average cannabis use frequency and small decrease in average cannabis misuse in the five years following recreational cannabis legalization in a community-based non-clinical cohort of Canadian adults. These changes were moderated by pre-legalization cannabis use, with more frequent pre-legalization consumers exhibiting the largest decreases in both outcomes. The apparent discrepancy between increasing cannabis use and decreasing cannabis misuse may have been driven by younger cannabis users, who typically transition from problematic to non-problematic use. This study also found that cannabis users’ product preferences evolved over the course of legalization away from dried flower and towards non-combustion products. Although longer-term follow-up is required, these results suggest a modest negative consequence (a small increase in cannabis use frequency) as well as some positive consequences (decrease in cannabis misuse and transition from combustible to non-combustible cannabis products) among adults following recreational cannabis legalization in Canada.

## Data Availability

All data produced in the present study are available upon reasonable request at the discretion of the authors

## Supplemental materials

**eTable 1:** Attrition analysis comparing those excluded due to having under 2 follow-ups to those included in the final study sample.

| Baseline characteristic | Included<br>(n=1,428) | Lost to Follow-up (n=51) | p-value |
| --- | --- | --- | --- |
| Sex |  |  | 0.148 |
| Female | 859 (60.2%) | 25 (49.0%) |  |
| Male | 569 (39.8%) | 26 (51.0%) |  |
| Age |  |  | 0.426 |
| Mean (SD) | 34.5 (13.9) | 36.1 (14.4) |  |
| Ethnicity |  |  | 0.151 |
| White | 1127 (78.9%) | 45 (88.2%) |  |
| Non-white | 301 (21.1%) | 6 (11.8%) |  |
| Marital status |  |  | 0.929 |
| Unmarried | 975 (68.3%) | 34 (66.7%) |  |
| Married | 453 (31.7%) | 17 (33.3%) |  |
| Household income |  |  | 0.011 |
| Less than \$45,000 | 442 (31.0%) | 17 (33.3%) | |
| \$45,000 to \$90,000 | 437 (30.6%) | 24 (47.1%) | |
| \$90,000+ | 549 (38.4%) | 10 (19.6%) | |
| Education |  |  | 0.109 |
| Post-secondary degree | 653 (45.7%) | 17 (33.3%) |  |
| No post-secondary degree | 775 (54.3%) | 34 (66.7%) |  |
| Cannabis use frequency (percentage of days using) |  |  | 0.0167 |
| Mean (SD) | 13.8 (30.1) | 27.8 (40.1) |  |
| CUDIT-R score |  |  | 0.0707 |
| Mean (SD) | 2.60 (4.13) | 3.98 (5.29) |  |

Notes: p-values calculated from chi-square and t-tests.

**eTable 2:** Sensitivity analysis repeating main analysis with multiple imputation via JointAI.

|  | Outcome 1:<br>Cannabis use<br>frequency |  | Outcome 2:<br>CUDIT-R |  |
| --- | --- | --- | --- | --- |
|  | *b* | *SE* | *b* | *SE* |
| *Main effect models* |  |  |  |  |
| Time (in years) | 0.34 | 0.08 | -0.08 | 0.01 |
| *Interaction models* |  |  |  |  |
| Time (in years) | 0.80 | 0.11 | 0.10 | 0.01 |
| Pre-legalization cannabis use |  |  |  |  |
| None | Ref | - | Ref | - |
| Less than monthly | 1.83 | 1.29 | 1.49 | 0.18 |
| Monthly | 9.06 | 1.50 | 3.11 | 0.22 |
| Weekly | 36.79 | 1.58 | 6.08 | 0.23 |
| Daily | 91.22 | 1.70 | 10.12 | 0.24 |
| Time* Pre-legalization<br>cannabis use |  |  |  |  |
| Time*Less than monthly | 0.43 | 0.22 | -0.21 | 0.03 |
| Time*Monthly | 0.61 | 0.25 | -0.30 | 0.03 |
| Time*Weekly | -1.01 | 0.28 | -0.48 | 0.04 |
| Time* Daily | -6.14 | 0.29 | -0.78 | 0.04 |
| Notes: All models adjusted for baseline age, sex, ethnicity, income, education, and marital status. |  |  |  |  |

**eTable 3:** Supplementary LMMs examining age and sex as moderators for change in cannabis use frequency and CUDIT-R in the 5 years following cannabis legalization.

|  | Cannabis use frequency |  |  |  | CUDIT-R |  |  |  |
| --- | --- | --- | --- | --- | --- | --- | --- | --- |
|  | *b* | *SE* | Omnibus test |  | *b* | *SE* | Omnibus test |  |
|  |  |  | *F* | p |  |  | *F* | p |
| Time (in years) | 0.29 | 0.14 | **18.56** | **<0.001** | -0.12 | 0.01 | **54.54** | **<0.001** |
| Baseline age (30+) | -0.20 | 1.11 | 0.03 | 0.858 | -0.55 | 0.16 | **12.37** | **<0.001** |
| Time*Baseline age | 0.11 | 0.16 | 0.49 | 0.486 | 0.09 | 0.02 | **19.39** | **<0.001** |
| Time (in years) | 0.44 | 0.10 | **15.03** | **<0.001** | -0.05 | 0.01 | **59.98** | **<0.001** |
| Sex (Male) | 0.97 | 0.96 | 1.03 | 0.311 | 0.51 | 0.13 | **14.75** | **<0.001** |
| Time*Sex | -0.25 | 0.16 | 2.37 | 0.124 | -0.06 | 0.02 | **7.74** | **<0.001** |

Notes: All models adjusted for baseline age, sex, ethnicity, income, education, and marital status. When testing age as a moderator, we dichotomized age to separate young adults, who typically age out of cannabis use,<sup>30</sup> from middle-aged and older adults.

**eFigure 1:**
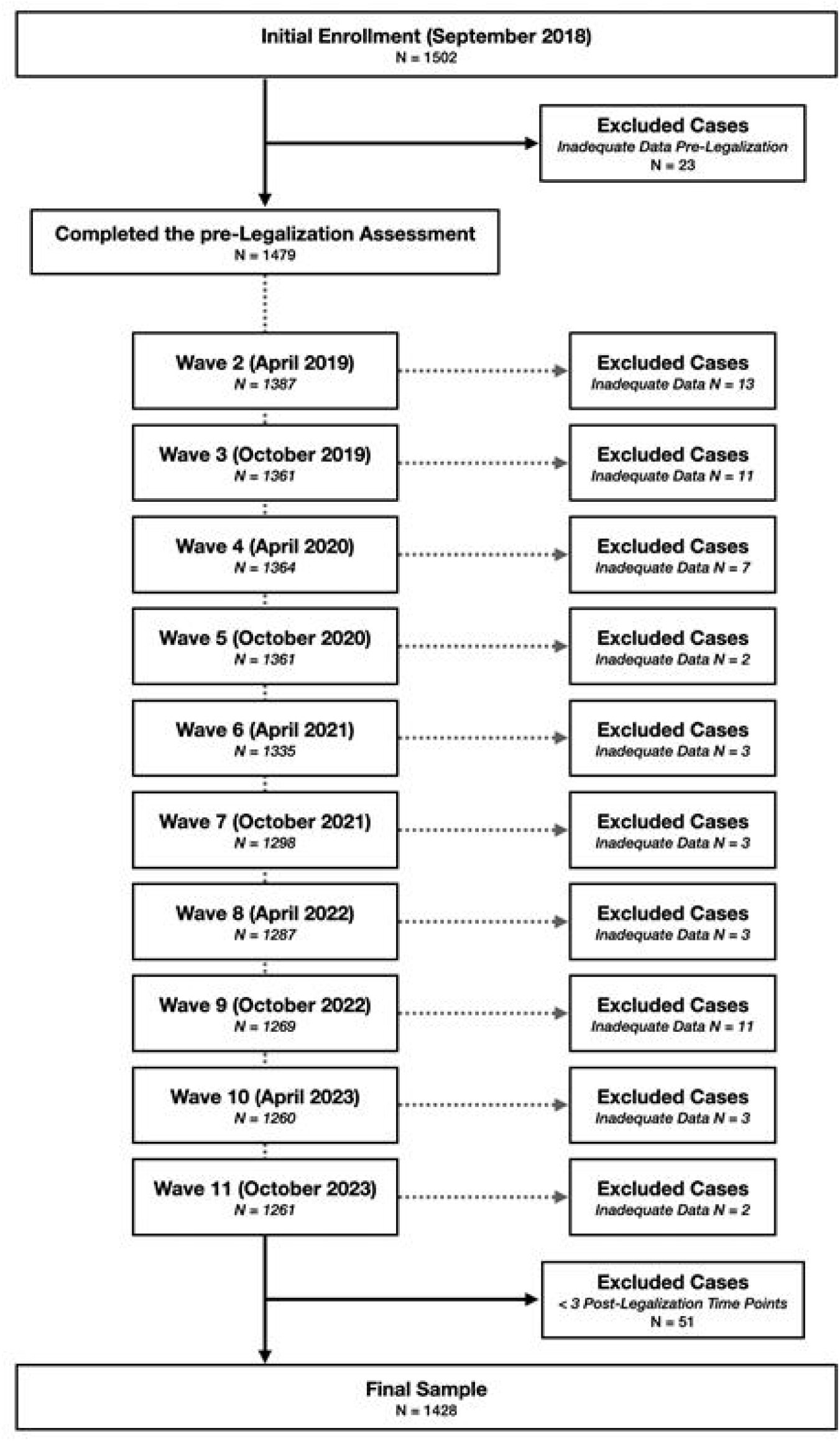
Flowchart of study cohort exclusions and loss to follow up by wave.

**eFigure 2:**
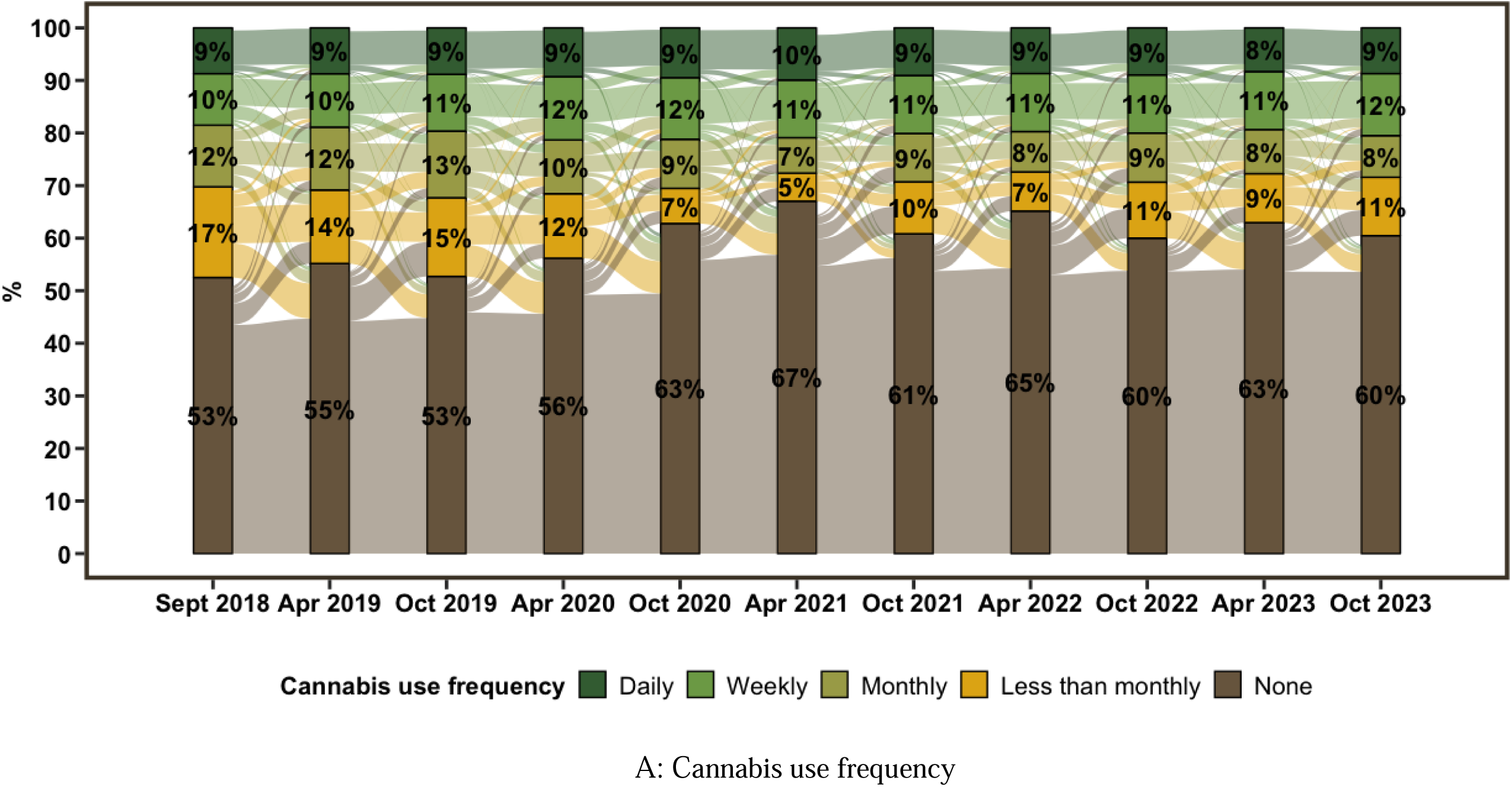
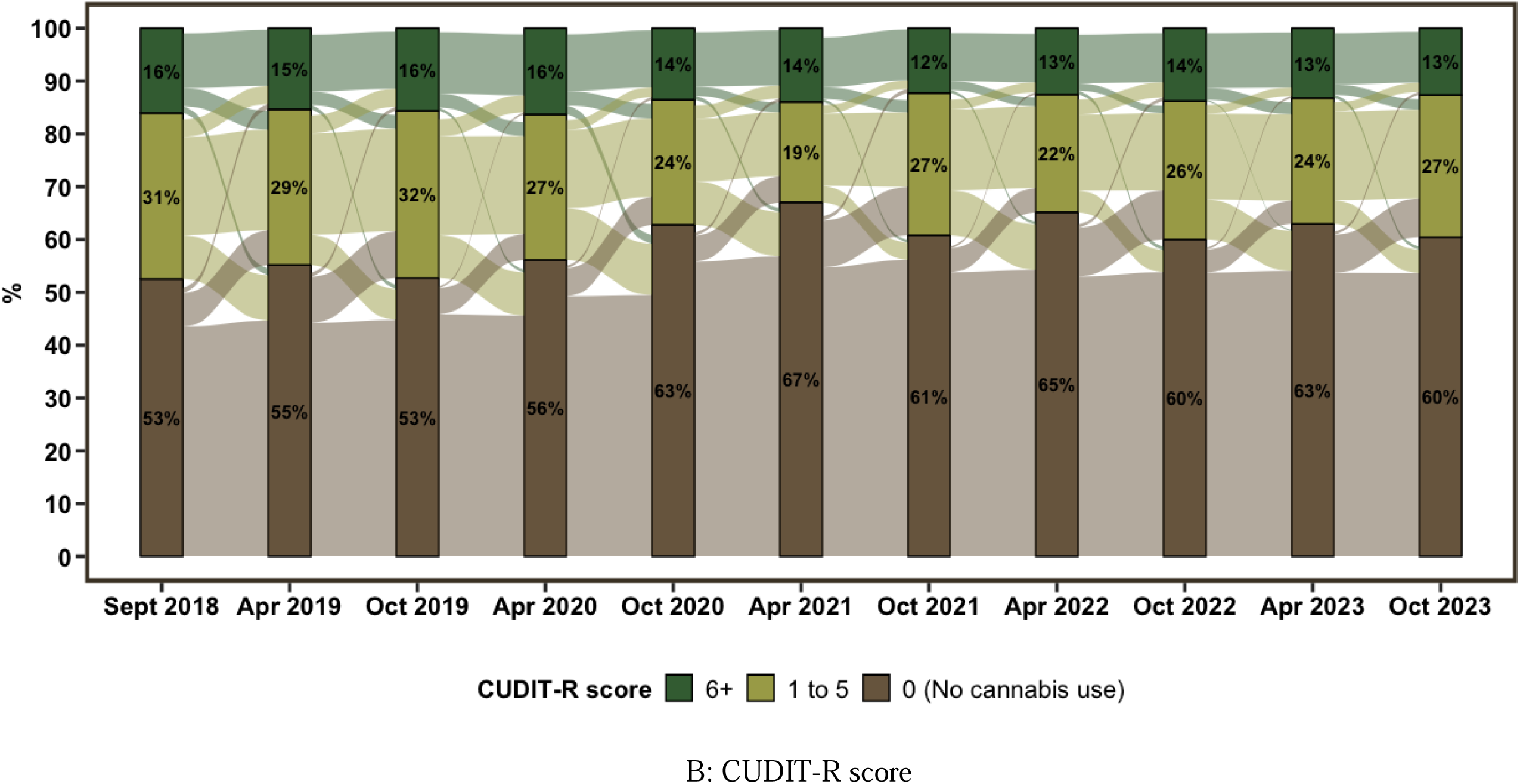
Alluvial plots showing transitions in cannabis use frequency and CUDIT-R score from pre-legalization (September 2018) to 5 years post-legalization (October 2023).

**eFigure 3:**
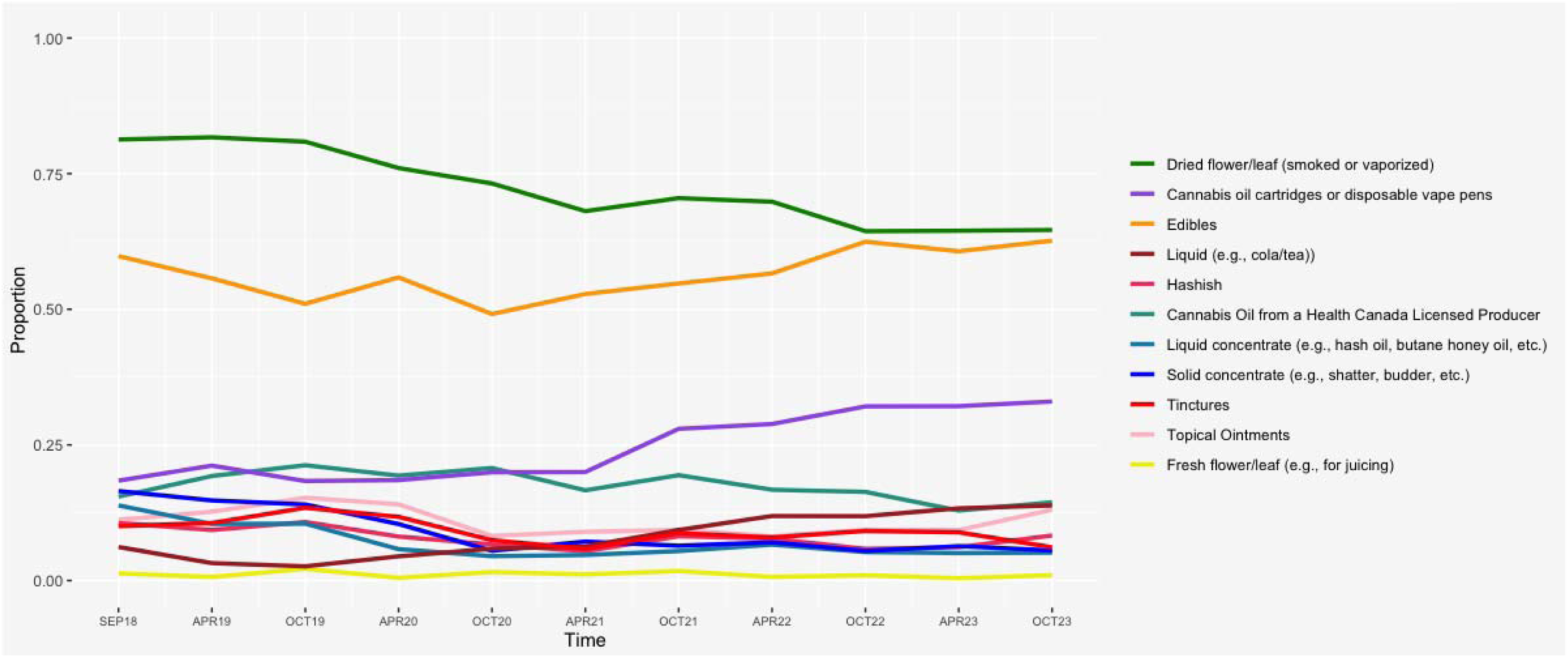
Cannabis product preferences over time since cannabis legalization among active cannabis users at each wave (all products)

**eFigure 4:**
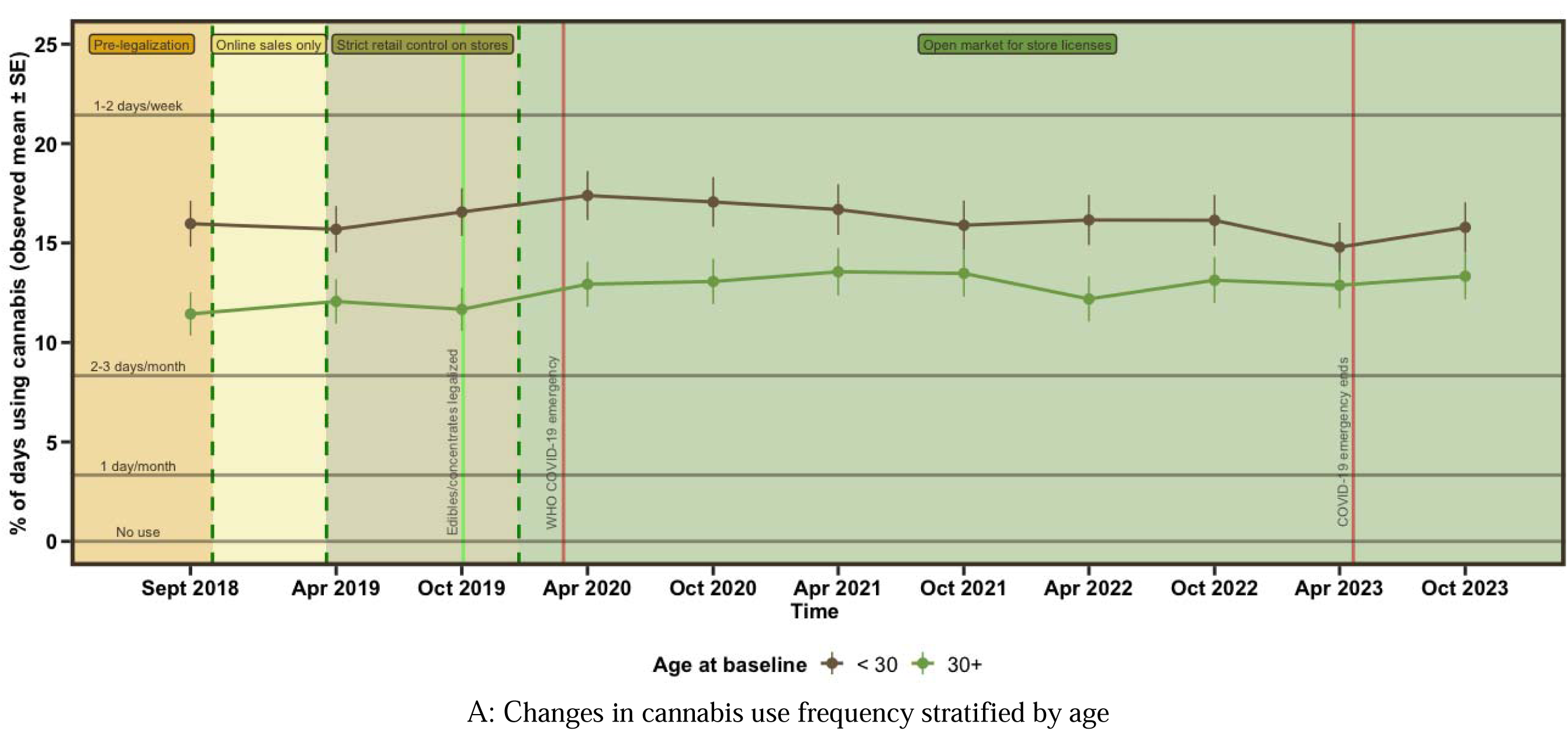
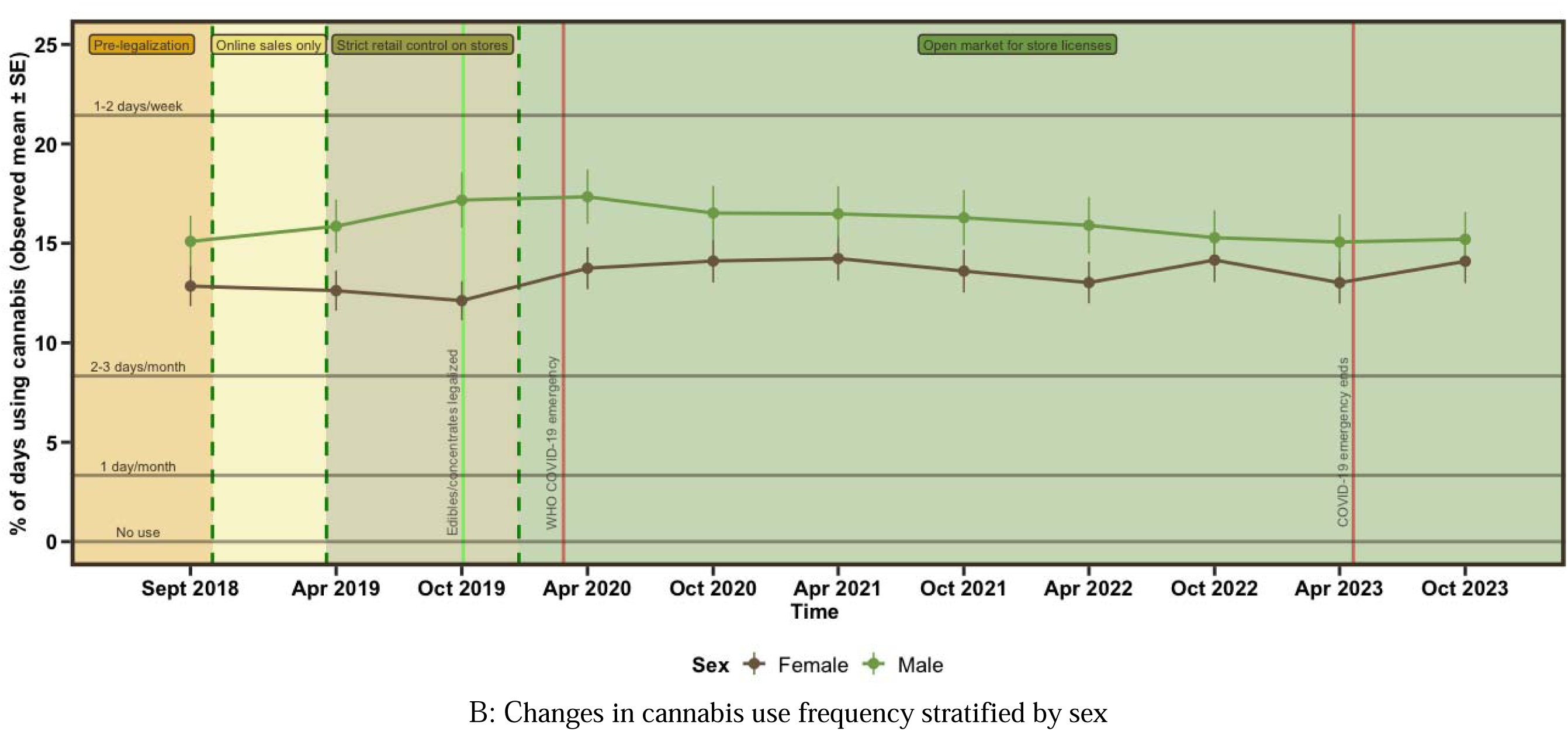
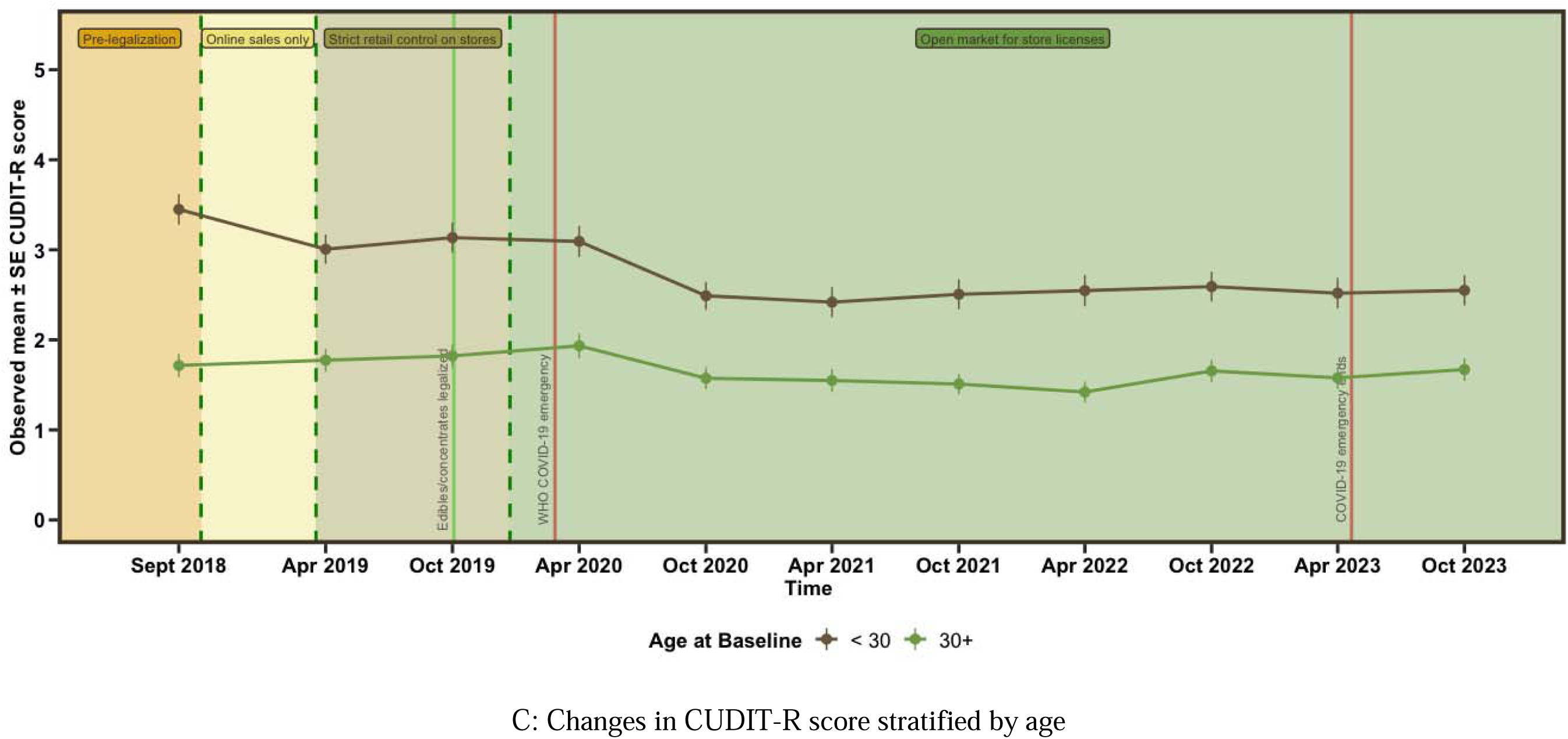
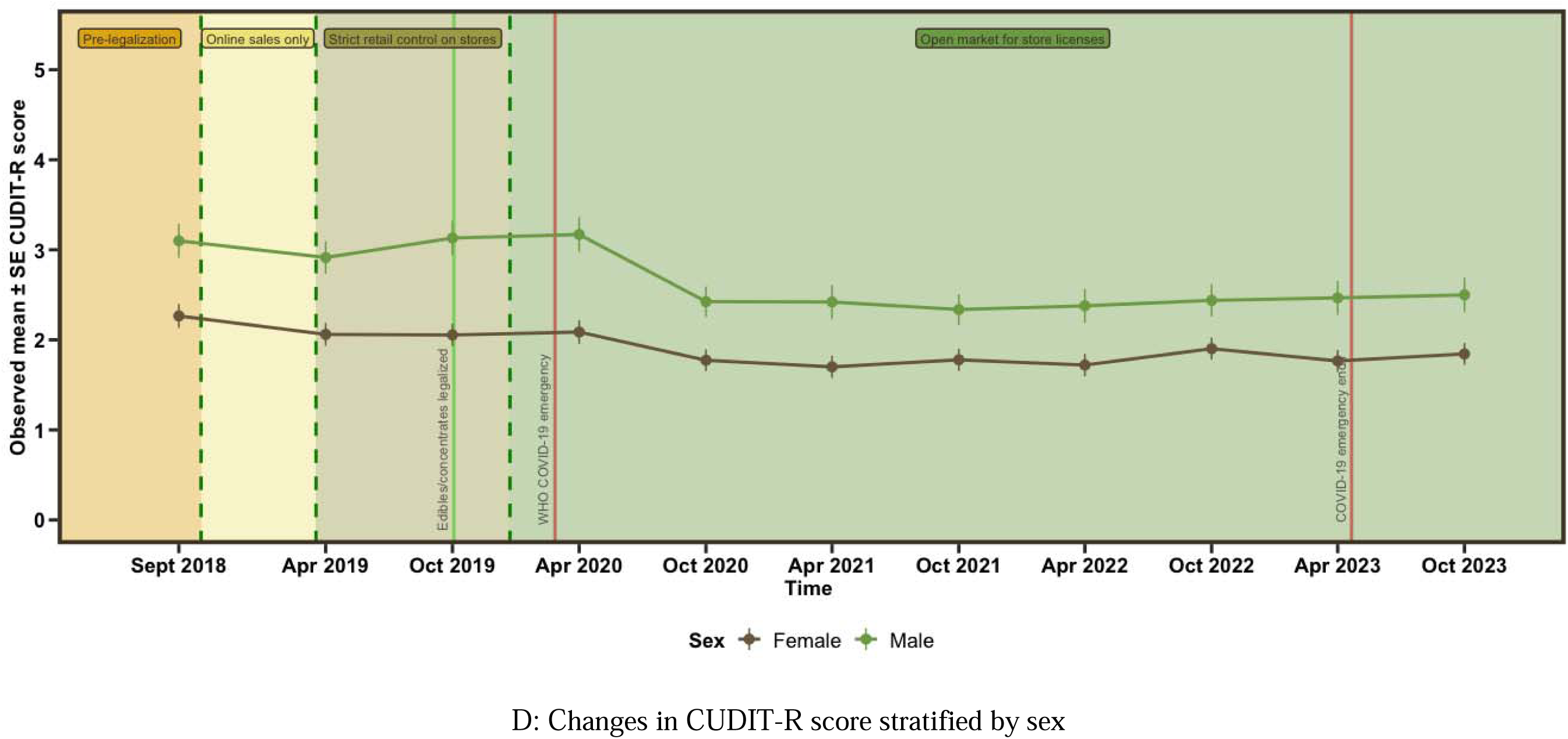
Mean (±SEM) for cannabis use frequency and CUDIT-R score over 5 years since legalization from September 2018 to October 2023 (10 waves) stratified by baseline age (subpanels A and C) and sex (subpanels B and D).

## References

1. Doggett A, Battista K, Jiang Y, de Groh M, Leatherdale ST. Patterns of Cannabis Use among Canadian Youth over Time; Examining Changes in Mode and Frequency Using Latent Transition Analysis. Subst Use Misuse. 2022;57(4):548–559. doi:10.1080/10826084.2021.2019785

2. Haines-Saah RJ, Fischer B. Youth Cannabis use and Legalization in Canada - Reconsidering the Fears, Myths and Facts Three Years In. J Can Acad Child Adolesc Psychiatry. 2021;30(3):191–196.

3. Hall W, Lynskey M. Evaluating the public health impacts of legalizing recreational cannabis use in the United States. Addiction. 2016;111(10):1764–1773. doi:10.1111/add.13428

4. McDonald AJ, Roerecke M, Mann RE. Adolescent cannabis use and risk of mental health problems-the need for newer data. Addiction. 2019;114(10):1889–1890. doi:10.1111/add.14724

5. Hammond D, Corsetti D, Goodman S, et al. International Cannabis Policy Study – Canada 2021 Summary.; 2022. https://cannabisproject.ca/wp-content/uploads/2022/10/2021-ICPS-National-Canada-Report-Sept-27.pdf

6. Mahamad S, Wadsworth E, Rynard V, Goodman S, Hammond D. Availability, retail price and potency of legal and illegal cannabis in Canada after recreational cannabis legalisation. Drug Alcohol Rev. 2020;39(4):337–346. doi:10.1111/dar.13069

7. Farrelly KN, Wardell JD, Marsden E, et al. The Impact of Recreational Cannabis Legalization on Cannabis Use and Associated Outcomes: A Systematic Review. Subst Abuse. 2023;17:11782218231172054. doi:10.1177/11782218231172054

8. Kerr WC, Lui C, Ye Y. Trends and age, period and cohort effects for marijuana use prevalence in the 1984 to 2015 US National Alcohol Surveys. Addiction. 2018;113(3):473–481. doi:10.1111/add.14031

9. Cerdá M, Mauro C, Hamilton A, et al. Association Between Recreational Marijuana Legalization in the United States and Changes in Marijuana Use and Cannabis Use Disorder From 2008 to 2016. JAMA Psychiatry. 2020;77(2):165–171. doi:10.1001/jamapsychiatry.2019.3254

10. Imtiaz S, Nigatu YT, Ali F, et al. Cannabis legalization and cannabis use, daily cannabis use and cannabis-related problems among adults in Ontario, Canada (2001–2019). Drug and Alcohol Dependence. 2023;244:109765. doi:10.1016/j.drugalcdep.2023.109765

11. Rotermann M. What has changed since cannabis was legalized? Health Rep. 2020;31(2):11–20. doi:10.25318/82-003-x202000200002-eng

12. Goodman S, Dann MJ, Fataar F, Abramovici H. How have cannabis use and related indicators changed since legalization of cannabis for non-medical purposes? Results of the Canadian Cannabis Survey 2018–2022. International Journal of Drug Policy. 2024;127:104385. doi:10.1016/j.drugpo.2024.104385

13. Myran DT, Tanuseputro P, Auger N, Konikoff L, Talarico R, Finkelstein Y. Pediatric Hospitalizations for Unintentional Cannabis Poisonings and All-Cause Poisonings Associated With Edible Cannabis Product Legalization and Sales in Canada. JAMA Health Forum. 2023;4(1):e225041. doi:10.1001/jamahealthforum.2022.5041

14. Myran DT, Pugliese M, Roberts RL, et al. Association between non-medical cannabis legalization and emergency department visits for cannabis-induced psychosis. Mol Psychiatry. Published online July 27, 2023:1–10. doi:10.1038/s41380-023-02185-x

15. Myran DT, Roberts R, Pugliese M, Taljaard M, Tanuseputro P, Pacula RL. Changes in Emergency Department Visits for Cannabis Hyperemesis Syndrome Following Recreational Cannabis Legalization and Subsequent Commercialization in Ontario, Canada. JAMA Network Open. 2022;5(9):e2231937. doi:10.1001/jamanetworkopen.2022.31937

16. Myran DT, Roberts R, Pugliese M, et al. Acute care related to cannabis use during pregnancy after the legalization of nonmedical cannabis in Ontario. CMAJ. 2023;195(20):E699–E708. doi:10.1503/cmaj.230045

17. Myran DT, Pugliese M, Tanuseputro P, Cantor N, Rhodes E, Taljaard M. The association between recreational cannabis legalization, commercialization and cannabis-attributable emergency department visits in Ontario, Canada: an interrupted time-series analysis. Addiction. 2022;117(7):1952–1960. doi:10.1111/add.15834

18. Callaghan RC, Sanches M, Vander Heiden J, Kish SJ. Impact of Canada’s cannabis legalisation on youth emergency department visits for cannabis-related disorders and poisoning in Ontario and Alberta, 2015–2019. Drug and Alcohol Review. 2023;42(5):1104–1113. doi:10.1111/dar.13637

19. Myran DT, Gaudreault A, Pugliese M, Tanuseputro P, Saunders N. Cannabis-involvement in emergency department visits for self-harm following medical and non-medical cannabis legalization. Journal of Affective Disorders. Published online February 1, 2024. doi:10.1016/j.jad.2024.01.264

20. Stall NM, Shi S, Malikov K, et al. Edible Cannabis Legalization and Cannabis Poisonings in Older Adults. JAMA Internal Medicine. Published online May 20, 2024. doi:10.1001/jamainternmed.2024.1331

21. Doggett A, Belisario K, McDonald AJ, Ferro MA, Murphy JG, MacKillop J. Cannabis Use Frequency and Cannabis-Related Consequences in High-Risk Young Adults Across Cannabis Legalization. JAMA Network Open. 2023;6(9):e2336035. doi:10.1001/jamanetworkopen.2023.36035

22. Turna J, Belisario K, Balodis I, Van Ameringen M, Busse J, MacKillop J. Cannabis use and misuse in the year following recreational cannabis legalization in Canada: A longitudinal observational cohort study of community adults in Ontario. Drug Alcohol Depend. 2021;225:108781. doi:10.1016/j.drugalcdep.2021.108781

23. Hall W, Stjepanović D, Dawson D, Leung J. The implementation and public health impacts of cannabis legalization in Canada: a systematic review. Addiction. n/a(n/a). doi:10.1111/add.16274

24. Fischer B, Lee A, Robinson T, Hall W. An overview of select cannabis use and supply indicators pre- and post-legalization in Canada. Subst Abuse Treat Prev Policy. 2021;16(1):77. doi:10.1186/s13011-021-00405-7

25. Schultz NR, Bassett DT, Messina BG, Correia CJ. Evaluation of the psychometric properties of the cannabis use disorders identification test - revised among college students. Addict Behav. 2019;95:11–15. doi:10.1016/j.addbeh.2019.02.016

26. Adamson SJ, Kay-Lambkin FJ, Baker AL, et al. An improved brief measure of cannabis misuse: the Cannabis Use Disorders Identification Test-Revised (CUDIT-R). Drug Alcohol Depend. 2010;110(1-2):137–143. doi:10.1016/j.drugalcdep.2010.02.017

27. Erler NS, Rizopoulos D, Lesaffre EMEH. JointAI: Joint Analysis and Imputation of Incomplete Data in R. Journal of Statistical Software. 2021;100(20):1–56. doi:10.18637/jss.v100.i20

28. Cheung YB. A modified least-squares regression approach to the estimation of risk difference. Am J Epidemiol. 2007;166(11):1337–1344. doi:10.1093/aje/kwm223

29. von Elm E, Altman DG, Egger M, et al. The Strengthening the Reporting of Observational Studies in Epidemiology (STROBE) statement: guidelines for reporting observational studies. J Clin Epidemiol. 2008;61(4):344–349. doi:10.1016/j.jclinepi.2007.11.008

30. Cristiano N, Sharif-Razi M. Marijuana, cocaine, and ecstasy in this day and age: A quantitative investigation of the “aging out” process. Journal of Substance Use. 2019;24(4):400–406. doi:10.1080/14659891.2019.1581288

31. Cousijn J, Kuhns L, Larsen H, Kroon E. For better or for worse? A pre–post exploration of the impact of the COVID 19 lockdown on cannabis users. Addiction. 2021;116(8):2104–2115. doi:10.1111/add.15387

32. Health Canada. Taking stock of progress: Cannabis legalization and regulation in Canada. Published September 22, 2022. https://www.canada.ca/en/health-canada/programs/engaging-cannabis-legalization-regulation-canada-taking-stock-progress/document.html

